# PREDICTORS OF UPTAKE OF A POTENTIAL COVID-19 VACCINE AMONG NIGERIAN ADULTS

**DOI:** 10.1101/2020.12.28.20248965

**Authors:** Charles Oluwatemitope Olomofe, Victor Kehinde Soyemi, Bolaji Felicia Udomah, Adeyinka Olabisi Owolabi, Emmanuel Eziashi Ajumuka, Chukwudum Martin Igbokwe, Uriel Oludare Ashaolu, Ayodele Olusola Adeyemi, Yetunde Bolatito Aremu-Kasumu, Olufunke Folasade Dada, John Chikezie Ochieze, Olaniyi Bamidele Fayemi, Kehinde Williams Ologunde, Gbenga Omotade Popoola, Olumuyiwa Elijah Ariyo

**Author notes:** Correspondence: Charles Oluwatemitope Olomofe.

## Abstract

**Background:** The Coronavirus diseases (COVID-19) pandemic is not abating and there is no approved treatment yet. The development of vaccines is hoped to help in addressing this disease outbreak. However, in the face of anti-vaccines uprise, it is important to understand the factors that may influence the uptake of COVID-19 vaccines as this will influence how successful the fight against COVID-19 will be in the long term.

**Methods:** A cross-sectional study among 776 adult Nigerians (age ≥18 years) was conducted in the 36 States of Nigeria and the Capital City with online questionnaire. The questionnaire consisted of 5 sections: socio-demographic characteristics of respondents, respondent’s knowledge of COVID-19, respondents risk perception of COVID-19, vaccination history of respondents, and willingness to receive COVID-19 vaccine. Descriptive analysis of variables was done and multivariate analysis using logistic regression was carried out to determine the predictors of uptake of a potential COVID-19 vaccine. The level of significance was predetermined at a p-value < 0.05. Data analysis was done with SPSS version 21.

**Results:** Most of the respondents were male (58.1%). Most participants were willing to take a potential COVID-19 vaccine (58.2%), while 19.2% would not take it with 22.6% indecisive. 53.5% would prefer a single dose COVID-19 vaccine. For vaccine uptake, being male (p= 0.002) and the perception that “vaccines are good” (p< 0.001) were the positive predictor of uptake of a potential COVID-19 vaccine.

**Conclusion:** Most Nigerians were willing to take a potential COVID-19 vaccine with the male gender and perception that “vaccines are good” being positive predictors. There is a need for public enlightenment aim at encouraging those that are indecisive or averse to receiving COVID-19 vaccines.

## BACKGROUND

Coronavirus disease 2019 (COVID-19), is caused by Severe Acute Respiratory Syndrome Corona virus-2 (SARS-CoV-2).^1^ As a result of the widespread occurrence and significant global health risk, the World Health Organization (WHO) declared COVID-19 a Public Health Emergency of International Concern (PHEIC) in late January 2020.^2^ Nigeria reported its index case on February 27, 2020, from an Italian citizen who tested positive in Lagos State, the commercial capital of the country.^3^ By the first week in March 2020, the number of cases had risen to over 100,000 affecting multiple continents, and WHO subsequently declared COVID-19 a global pandemic on March 11, 2020.^4^

As of December 4, 2020, there were over 65 Million cases of COVID-19 across the globe and more than 1.5 million deaths. In Nigeria, data showed over 68,000 cases with about 1200 deaths^5^. Despite the devastating scourge of the disease, there has not been any proven treatment against the SARS-CoV-2. The recommendations of the disease control bodies are focused on disease prevention and control measures to minimize spread and to reduce the burden on the healthcare system.^6-8^ Vaccination is an effective preventive measure reducing morbidity and mortality caused by infectious agents. It constitutes the mainstay of prevention of infectious childhood diseases and is of major importance in primary health care.^9^ Since the onset of the COVID-19 pandemic, there has been a significant drive to find an effective vaccine with several vaccines at various stages of drug trials and some already being tested in humans across various countries.^10,11^ However, the success of this vaccine will be dependent on the acceptance and uptake level and the subsequent development of herd immunity.^9^ In recent times, there has been a growing anti-vaccine movement,^11^ it is therefore imperative to explore the barriers and drivers of uptake of a potential COVID-19 vaccine to help the government, policymakers, and health care workers mitigate the impact of probable low vaccine uptake.

Vaccine uptake refers to the absolute number of people who receive a specified vaccine dose(s);^12^ and low uptake has been increasingly recognized as a challenge to the success of vaccination programs.^11^ Uptake of vaccines can be influenced by several factors such as personal risk perception, fear of side effects, access to media, information sources, religious/cultural beliefs, the convenience of getting to a health facility, level of trust for the healthcare system, household wealth, residence, ethnicity, and other demographic variables, as well as other social influences.^13,14^ Introduction of a new vaccine to the public may be met with hesitancy due to skepticism about its effectiveness and potential safety.^9^ Several studies have been carried out on determinants of uptake of already existent vaccines^14-16^ but there is currently a paucity of data on the uptake of a potential COVID-19 vaccine.

Adults are thought to be the important drivers of SARS-CoV-2 transmission in the community and are also known to be more susceptible than children. Unfortunately, vaccine uptake among adults in Nigeria is already poor.^17,18^ There is, therefore, a concern as to the likely picture of uptake of a potential COVID-19 vaccine in the adult population. This study, therefore, aimed to investigate the willingness to receive the COVID-19 vaccine among the Nigerian adult population and the predictors of uptake of the vaccine.

## METHODS

This was a cross-sectional study carried out in Nigeria, the most populous nation in Africa with an estimated population of about 200 million and a total land area of 910,770 Km2 (351,650 sq. miles).^19^ Nigeria has six geopolitical zones (Southwest, Southeast, Southsouth, Northwest, Northeast, Northcentral) with 36 states. Ethical approval was obtained from the Health Research Ethics Committee of the Federal Medical Centre Gusau, Zamfara State, Nigeria.

The study was conducted online using a pre-tested, semi-structured questionnaire and included Nigerian adults above the age of 18 years who consented to participate in the study. The minimum sample size was determined to be 409 at a confidence level of 95% and based on the proportion of people with good knowledge of 39% in a previous study^20^ and a 5% margin of error.

The questionnaire was adapted from several published literature^20-23^ and comprised of five sections A-E namely, Socio-demographic characteristics of respondents, respondent’s knowledge of COVID-19, respondents risk perception of COVID-19, vaccination history of respondents, and willingness to receive COVID-19 vaccine. The pre-test of the questionnaire was done on 10% of the subjects each at six different states from the six geopolitical zones and was not included in the study. The pretested questionnaires with participants’ information sheets were distributed widely online. The data were collected from June to July 2020.

Data were analyzed using SPSS version 21. The knowledge of COVID-19 among respondents was scored based on the number of correct responses given. The number of correct responses was compared with the average score. Participants whose score equaled or was above the average were categorized as having good knowledge while those who scored below the average were categorized as having poor knowledge. Descriptive statistics (frequency tables and percentages) were calculated for the sample demographic characteristics. The frequency and percentage of willingness to receive the COVID-19 vaccine were also calculated. Chi-square analysis was computed to test for association between sociodemographic characteristics, knowledge of COVID-19 among respondents, vaccination history of the respondent, and willingness to receive a COVID-19 vaccine. Multivariate analysis using logistic regression was carried out to determine the predictors of uptake of a potential COVID-19 vaccine. The level of significance was predetermined at a p-value of less than 0.05 at a 95% confidence level.

## RESULTS

A total of 776 participants completed the survey. Most were within the ages of 36-45 years (43.9%), with 58.1% males and 40.9% females. The majority (53.2%) have tertiary education with 7.7% of the respondents being artisans, 8.6% being teachers and 25% being health care workers. Most respondents (26.7%) preferred not to say their annual income with 17.5% earning less than 500,000 Naira /Annum. Zonal representation of respondents (State of origin and place of residence) revealed that most of the respondents were from the southwest zone of the country (47.4% and 46% respectively). Most respondents’ households are made up of 1-4 persons (48.3%) while 5.7% of the respondents have more than 8 persons per household (**Table 1**).

**Table 1:** Socio-demographic characteristics of Respondents.

| **Variables** | **Frequency (*n* = 776)** | **%** |
| --- | --- | --- |
| **Age group (years)** |  |  |
| *18 – 25* | 93 | 12.0 |
| *26 – 35* | 241 | 31.1 |
| *36 – 45* | 341 | 43.9 |
| *46 – 55* | 73 | 9.4 |
| *> 55* | 28 | 3.6 |
| **Gender** |  |  |
| *Male* | 451 | 58.1 |
| *Female* | 317 | 40.9 |
| *Prefer not to say* | 8 | 1.0 |
| **Marital Status** |  |  |
| *Single* | 214 | 27.6 |
| *Married* | 489 | 63.0 |
| *Separated/ Divorced* | 15 | 1.9 |
| *Widowed/ Widower* | 7 | 0.9 |
| *In a relationship* | 45 | 5.8 |
| *Undisclosed* | 6 | 0.8 |
| **Religion** |  |  |
| *None* | 9 | 1.2 |
| *Christianity* | 616 | 79.4 |
| *Islam* | 147 | 18.9 |
| *Others* | 4 | 0.5 |
| **Level of Education** |  |  |
| *Secondary school or less* | 36 | 4.6 |
| *Tertiary* | 413 | 53.2 |
| *Postgraduate* | 327 | 42.1 |
| **Occupation** |  |  |
| *Unemployed* | 60 | 7.7 |
| *Trader* | 22 | 2.8 |
| *Teacher* | 67 | 8.6 |
| *Artisan* | 17 | 2.2 |
| *Healthcare worker* | 194 | 25.0 |
| *Information technology* | 43 | 5.5 |
| *Financial services* | 45 | 5.8 |
| *Security agent* | 12 | 1.5 |
| *Agricultural services* | 19 | 2.4 |
| *Others* | 297 | 38.3 |
| **Estimated Income of Caregiver**<br>**Per Annum (Naira)** |  |  |
| *< 500,000* | 136 | 17.5 |
| *500,000 -1 million* | 120 | 15.5 |
| *1 – 2 million* | 117 | 15.1 |
| *2 – 3 million* | 54 | 7.0 |
| *3 – 4 million* | 37 | 4.8 |
| *> 4 million* | 105 | 13.5 |
| *I prefer not to say* | 207 | 26.7 |
| **State Origin (Zone)** |  |  |
| *North-Central* | 90 | 11.6 |
| *North-East* | 46 | 5.9 |
| *North-West* | 49 | 6.3 |
| *South-East* | 119 | 15.3 |
| *South-South* | 94 | 12.1 |
| *South-West* | 368 | 47.4 |
| *Prefer not to say* | 10 | 1.3 |
| **Place of residence (Zone)** |  |  |
| *North-Central* | 174 | 22.4 |
| *North-East* | 24 | 3.1 |
| *North-West* | 103 | 13.3 |
| *South-East* | 32 | 4.1 |
| *South-South* | 40 | 5.2 |
| *South-West* | 357 | 46.0 |
| *Prefer not to say* | 46 | 5.9 |
| **No person in a household** |  |  |
| *1 – 4* | 375 | 48.3 |
| *5 – 8* | 357 | 46.0 |
| *> 8* | 44 | 5.7 |

Most of the respondents (58.2%) were willing to receive the vaccine once it is available while 19.2% of the respondents were not willing to receive the vaccine; 22.6% of the respondents were indecisive (**Table 2**). Many of the respondents who were unwilling to take the vaccine were not sure of reasons why they were unwilling to receive the vaccine (**Table 3**). Among the respondents that were willing to take the vaccine, the majority (53.5%) were comfortable with a single dose of the vaccine, while only 8% of the respondents were willing to take more than 4 doses. (**Table 4**) The preferred route of administration among most of the respondents was either oral (58.2%) or Injection (53.8%) with many of the respondents rejecting the administration of the vaccine through the intranasal route (76.1%) **(Table 5**).

**Table 2:** Respondents willingness to take potential COVID-19 Vaccine.

| **Willingness to take potential COVID**<br>**19 Vaccine** | **Frequency (*n* = 776)** | **Percent (%)** |
| --- | --- | --- |
| *Yes* | 452 | 58.2 |
| *No* | 149 | 19.2 |
| *Maybe* | 175 | 22.6 |

**Table 3:** Respondents reasons for refusing to take a Potential COVID-19 Vaccine.

|  | **Yes** | **No** | **Maybe** | **Don't** |
| --- | --- | --- | --- | --- |

| **Determinants** | **n (%)** | **n (%)** | **n (%)** | **know**<br>**n (%)** |
| --- | --- | --- | --- | --- |
| *The potential vaccine is a mark of the beast* | 4 (2.7) | 29 (19.5) | 9 (6.0) | 107 (71.8) |
| *The potential vaccine is to reduce the world population* | 4 (2.7) | 20 (13.4) | 20 (13.4) | 105 (70.5) |
| *The potential vaccine can make one sick* | 7 (4.7) | 10 (6.7) | 25 (16.8) | 107 (71.8) |
| *I might not be able to afford the vaccine* | 9 (6.0) | 21 (14.1) | 15 (10.1) | 104 (69.8) |
| *I don't have time to take any vaccine* | 3 (2.0) | 35 (23.5) | 8 (5.4) | 103 (69.1) |
| *My religion does not allow vaccination* | 0 (0.0) | 45 (30.2) | 4 (2.7) | 100 (67.1) |
| *I will take it if I get paid for it* | 5 (3.4) | 30 (20.1) | 15 (10.1) | 99 (66.4) |
| *I have other medical condition that would not allow me to take it* | 3 (2.0) | 38 (25.5) | 7 (4.7) | 101 (67.8) |
| *The vaccines we are taking are already too much* | 8 (5.4) | 26 (17.4) | 11 (7.4) | 104 (69.8) |
| *Vaccines are only for children* | 0 (0.0) | 44 (29.5) | 5 (3.4) | 100 (67.1) |
| *The vaccine is not needed because the infection is harmless* | 0 (0.0) | 40 (26.8) | 8 (5.4) | 101 (67.8) |

**Table 4:** Respondents preferred maximum dose(s) of potential COVID-19 vaccine.

| **Number of willing Permissible Dose** | **Frequency (*n* = 452)** | **Percent (%)** |
| --- | --- | --- |
| *1* | 242 | 53.5 |
| *2* | 116 | 25.7 |
| *3* | 50 | 11.1 |
| *≥ 4* | 36 | 8.0 |
| *Indecisive* | 8 | 1.8 |

**Table 5:** Respondents preferred route for potential COVID-19 vaccine.

| **The preferred route of administration** | **Yes**<br>**n (%)** | **No**<br>**n (%)** | **Maybe**<br>**n (%)** |
| --- | --- | --- | --- |
| *Oral* | 263 (58.2) | 170 (37.6) | 19 (4.2) |
| *Intranasal* | 62 (13.7) | 344 (76.1) | 46 (10.2) |
| *Injection* | 243 (53.8) | 174 (38.5) | 35 (7.7) |
| *Any Route* | 58 (12.8) | 336 (74.3) | 58 (12.8) |

Socio-demographic variables such as gender and religion of the respondents showed a statistically significant association with willingness to receive the vaccine (**Table 6a and 6b**). ‘Perception that vaccines are good or bad’, previous history of vaccination, and knowledge of COVID-19 were also shown to have a statistically significant association with willingness to receive the vaccine. However, there was no statistically significant association between risk perception of COVID-19 and willingness to receive COVID-19 vaccine (Table 6b). Male gender and ‘perception that vaccines generally are good’ were found to be significant predictors of uptake of a potential COVID-19 vaccine. Males were one and a half times more likely to receive the vaccine than females and those with a ‘perception that vaccines are generally good’ were seven times more likely to receive the vaccine than those who have a ‘perception that vaccines are generally bad’ (**Table 7)**.

**Table 6a:**
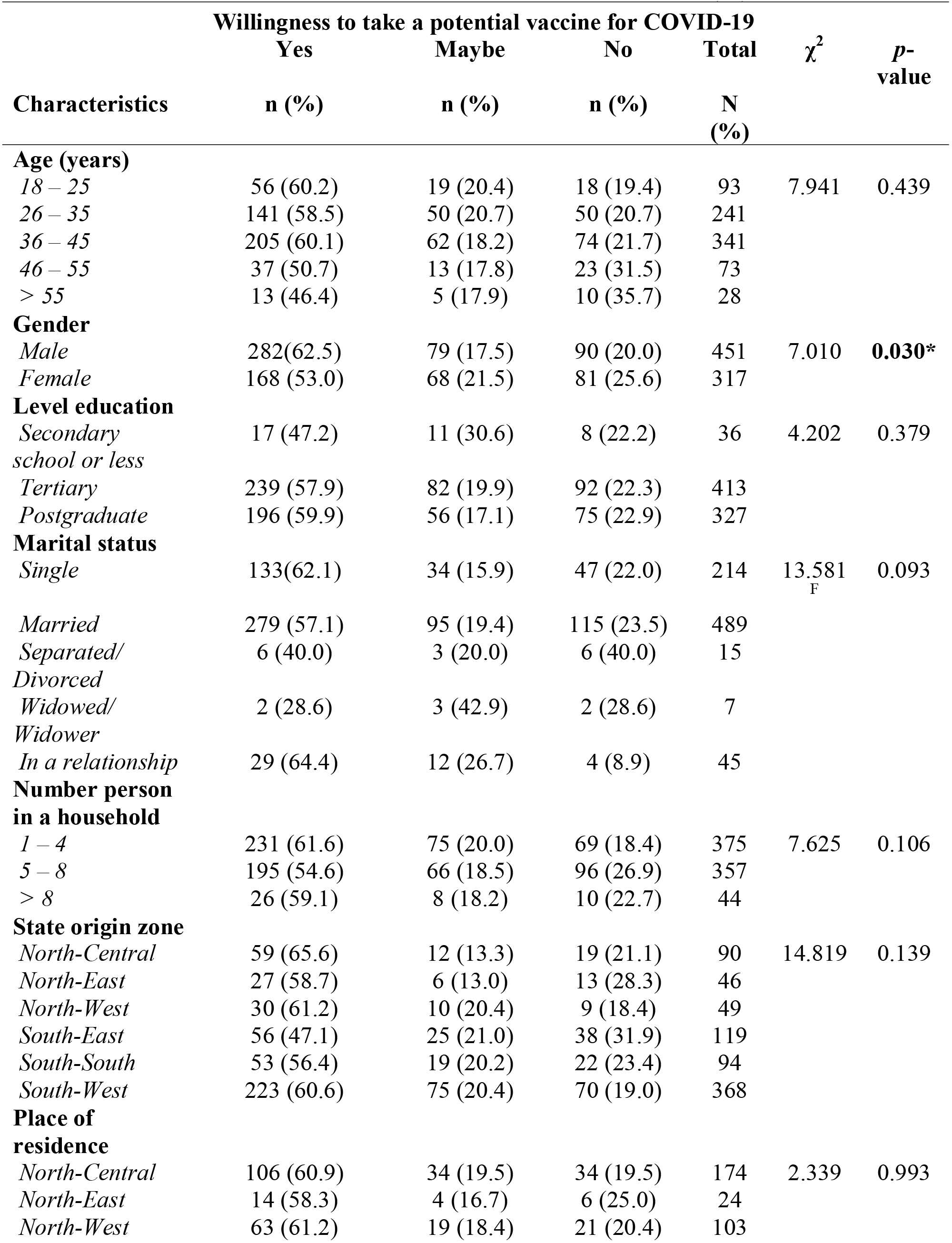
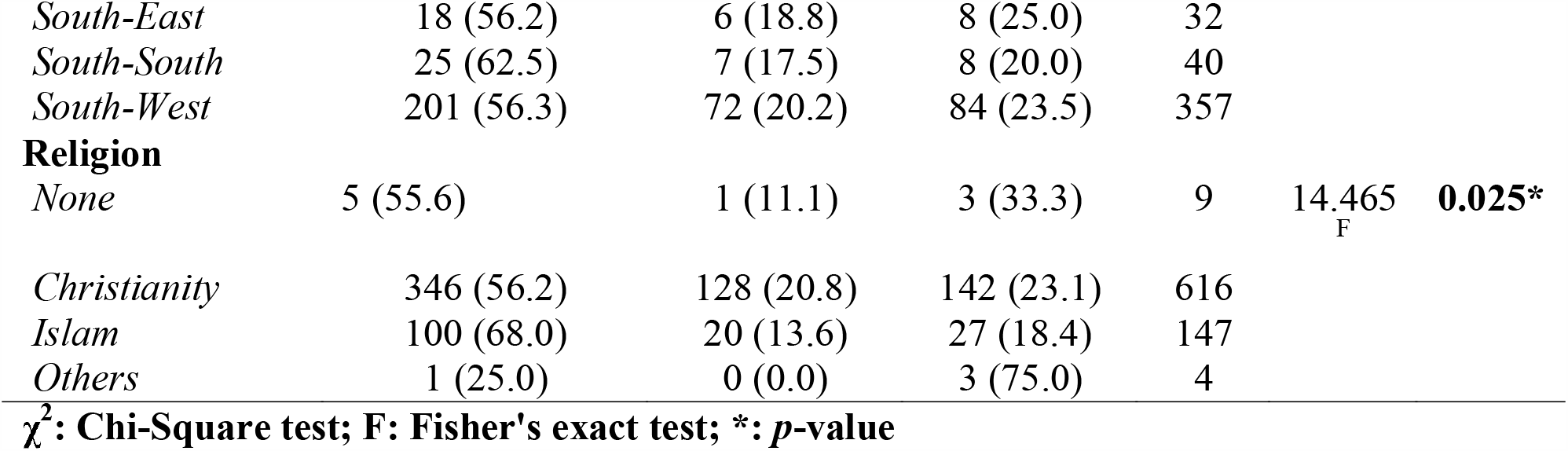
Willingness to be vaccinated by Respondents characteristics.

**Table 6b:** Willingness to be vaccinated by Respondents characteristics.

| Characteristics | Readiness to take a potential vaccine for COVID-19 | | | | $\chi^2$ | *p*-value |
| --- | --- | --- | --- | --- | --- | --- |
|  | Yes<br>n (%) | Maybe<br>n (%) | No<br>n (%) | Total<br>N (%) |  |  |
| **Occupation** |  |  |  |  |  |  |
| *Unemployed* | 35 (58.3) | 16 (26.7) | 9 (15.0) | 60 | 28.694 | **0.050** |
| *Trader* | 17 (77.3) | 2 (9.1) | 3 (13.6) | 22 |  |  |
| *Teacher* | 30 (44.8) | 17 (25.4) | 20 (29.9) | 67 |  |  |
| *Artisan* | 13 (76.5) | 0 (0.0) | 4 (23.5) | 17 |  |  |
| *Healthcare worker* | 124 (63.9) | 31(16.0) | 39 (20.1) | 194 |  |  |
| *Information technology* | 19 (44.2) | 9 (20.9) | 15 (34.9) | 43 |  |  |
| *Financial services* | 24 (53.3) | 10 (22.2) | 11 (24.4) | 45 |  |  |
| *Security agent* | 9 (75.0) | 1 (8.3) | 2 (16.7) | 12 |  |  |
| *Agricultural services* | 14 (73.7) | 4 (21.1) | 1 (5.3) | 19 |  |  |
| *Others* | 167 (56.2) | 59 (19.9) | 71 (23.9) | 297 |  |  |
| **Average income** |  |  |  |  |  |  |
| *< 500,000* | 85 (62.5) | 21(15.4) | 30 (22.1) | 136 | 9.780 | 0.460 |
| *500,000 -1 million* | 62 (51.7) | 25(20.8) | 33 (27.5) | 120 |  |  |
| *1 – 2 million* | 73 (62.4) | 20 (17.1) | 24 (20.5) | 117 |  |  |
| *2 – 3 million* | 37 (68.5) | 9 (16.7) | 8 (14.8) | 54 |  |  |
| *3 – 4 million* | 24 (64.9) | 9 (24.3) | 4 (10.8) | 37 |  |  |
| *> 4 million* | 61 (58.1) | 19 (18.1) | 25 (23.8) | 105 |  |  |
| **Vaccines (immunization) are good** |  |  |  |  |  |  |
| *Yes* | 419 (68.1) | 98 (15.9) | 98 (15.9) | 615 | 123.715 | **<0.001**<br>* |
| *No* | 33 (20.5) | 51(31.7) | 77 (47.8) | 11 |  |  |
| **Ever received any vaccine before** |  |  |  |  |  |  |
| *Yes* | 415 (59.5) | 136 (19.5) | 147 (21.1) | 698 | 8.879 | **0.012\*** |
| *No* | 37 (47.4) | 13 (16.7) | 28 (35.9) | 78 |  |  |
| **Risk perception for COVID-19** |  |  |  |  |  |  |
| *Good* | 289 (59.0) | 93 (19.0) | 108 (22.0) | 490 | 0.310 | 0.856 |
| *Poor* | 163 (57.0) | 56 (19.6) | 67 (23.4) | 286 |  |  |
| **Knowledge of COVID-19** |  |  |  |  |  |  |
| *Good* | 419 (59.8) | 135 (19.3) | 147 (21.0) | 701 | 10.950 | <0.001* |
| *Poor* | 33 (44.0) | 14 (18.7) | 28 (37.3) | 75 |  |  |

$\chi^2$ : Chi square test; \*: $p$ value <0.05

**Table 7:**
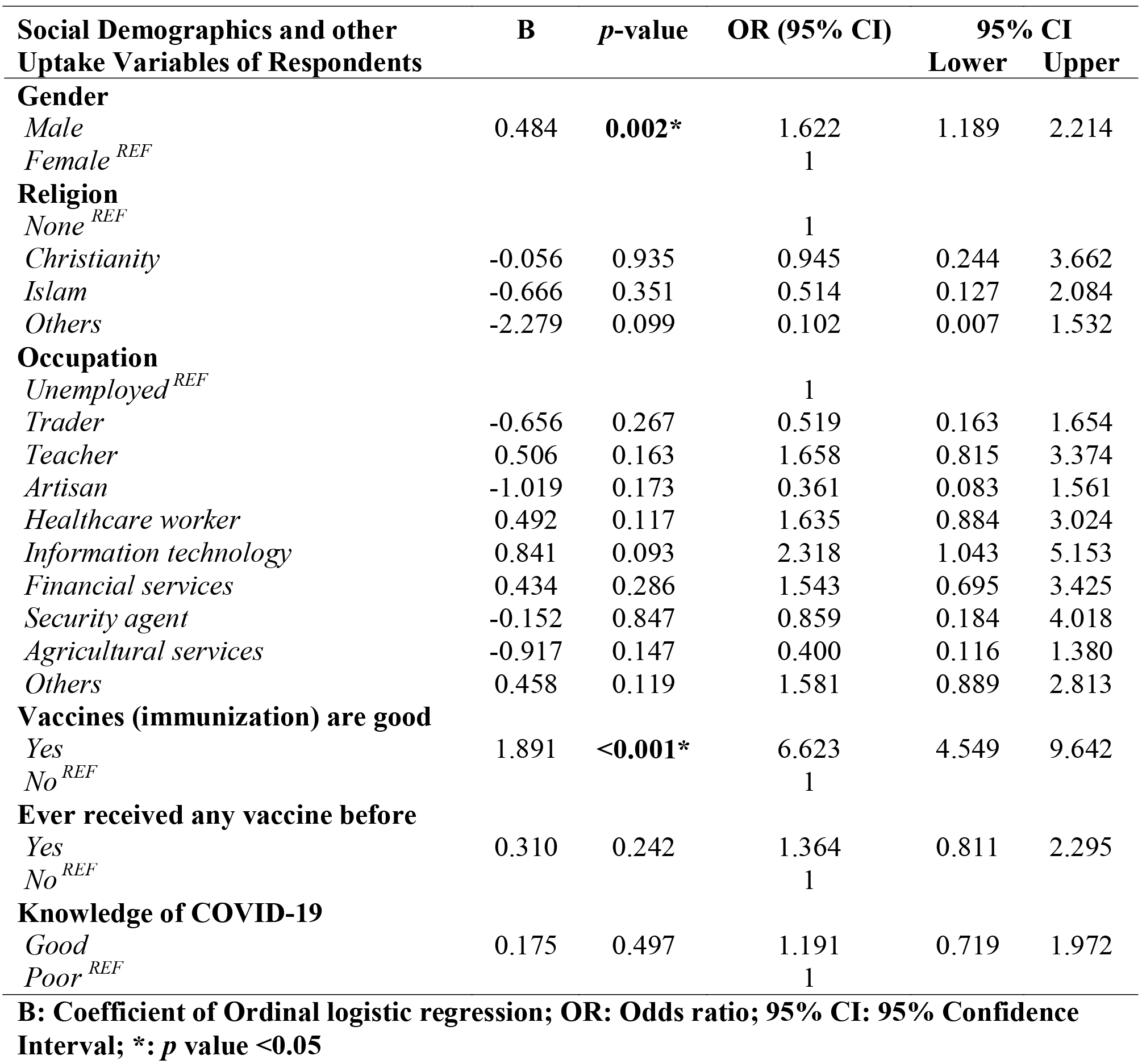
Predictors of uptake of a potential COVID-19 vaccine.

## DISCUSSION

This study reveals that only a simple majority (58.2%) were willing to take a COVID 19 vaccine when available. This is short of the findings from a study by Abdelhafiz A. et.al^24^ where about 73% of the respondents were willing to take a potential COVID-19 vaccine. Malik A.^25^ in his study in the United States (US) also found that 67% of participants would be willing to accept the vaccine. About one-fifth of the respondents in this study were indecisive; a finding similar to the one reported among healthcare workers from a study conducted by Fu C. et.al.^26^ Strikingly, there was an aversion to multiple vaccine doses among those willing to take the vaccine with most preferring the oral and the injection routes over other potential routes of administration This information is important for policymakers to improve uptake and compliance, especially if multiple route options become available. About one-fifth of the respondents were unwilling to receive a potential vaccine which appears to stem from skepticism about the safety of the potential vaccine. For instance, in this study, the majority were unsure if the potential vaccine is ‘a mark of the beast’ or if the motive is ‘to reduce the world population’. This is comparable to the findings in the study by Thunstrom L. et.al^9^ where it was reported that 20% of the US population would decline the vaccine. They noted that distrust of vaccine safety and vaccine novelty are among the most important obstacles to vaccination.

Five factors namely, gender, religion, occupation, knowledge of COVID-19, ‘perception that vaccines generally are good’, and previous vaccination(s) were shown to have a statistically significant association with ‘willingness to take vaccine’. However, male gender and ‘perception that vaccines generally are good’ were found to be the only independent significant predictors of uptake of a potential COVID 19 vaccine. Like this, Wang J.^27^ in his study in China found that being male increased the probability of accepting the vaccine. Contrary to this Padhi^28^ in his study in Saudi Arabia reported no association between gender and willingness to take a COVID-19 vaccine while Lazarus J ^29^ also reported that gender differences were small in his study across 19 Countries.

While gender (male), and ‘perception that vaccines generally are good’ predicted the positive uptake of a potential COVID-19 vaccine as hypothesized, surprisingly, knowledge and risk perception of the disease did not, despite reported good knowledge and positive risk perception. This suggests there are still other factors influencing the uptake of a potential vaccine; of note is the range of misconceptions bordering on the skepticism about the safety of a potential vaccine identified in this study, which seems to agree with the study by Thunstrom L. et.al.^9^ While it is recognized that hypothetical choices may not always reflect real-life behaviour/decision, it is imperative for stakeholders (for example government agencies/policymakers, non-governmental organizations, and health care workers) to still do more in terms of health education and promotion especially in addressing these misconceptions about a potential COVID-19 vaccine. This may go a long way in improving probable low vaccine uptake.

### Limitations

Findings may be influenced by selection bias because respondents needed access to a smartphone or computer. This may have excluded the poor, elderly who are most vulnerable to COVID-19 this may limit external validity and may have distorted estimation of those willing to take the vaccine.

## CONCLUSION

Our study showed that 58.2%of our sample from across Nigeria would be willing to take a COVID-19 vaccine. Male gender was the most significant independent predictor of vaccine uptake, this is particularly important in Nigeria where the patriarchal system is dominant. Most respondents who were not willing to receive vaccines could not give a reason for their stance. Policymakers and Stakeholders will need to focus attention on Health education campaigns targeted at both males and females in the community to improve the acceptance and uptake of the vaccine.

## Data Availability

The datasets used and/or analyzed during the current study are available from the corresponding author on reasonable request

## DECLARATIONS

### Ethics approval

Ethical approval was obtained from the Health Research Ethics Committee of Federal Medical Center, Gusau, Zamfara State, Nigeria

### Consent for publication

Not applicable

## Authors’Contributions

Conception/design of the study-COO, VKS, OBF, AOO; data collection-UOA, CMI, JCO, OFA, KWO; data analysis and interpretation-EEA, GOP, AOA, OEA; article drafting-BFU, YBA, OFA, AOO; Critical revision of the article-COO, BFU, VKS; final approval of the version to be published-all authors

## Acknowledgment

Ruth Oluwafunmike Olomofe is appreciated for proof-reading the final manuscript.

## REFERENCES

1. Baron S. Galveston; Structure and classification of viruses. Medical microbiology 4th edition.

2. Euro surveillance Editorial Team. Note from the editors: World Health Organization declares novel coronavirus (2019-nCoV) sixth public health emergency of international concern. Euro Surveill. 2020; 25 (5):200131e.

3. Nigeria Centre for Disease Control (NCDC). Nigeria records its first case of coronavirus disease (2020). Retrieved from https://ncdc.gov.ng/news/227/first-case-of-corona-virus-disease-confirmed-in-nigeria. Retrieved March 19th, 2020. [Online Resource]

4. Shigemura, J., Ursano, R. J., Morganstein, J. C., et al. (2020). Public responses to the novel 2019 coronavirus (2019-nCoV) in Japan: Mental health consequences and target populations. Psychiatry and Clinical Neurosciences. https://doi.org/10.1111/pcn.12988

5. COVID-19 situation update worldwide as of 4 December 2020. [Internet]. 2020 [cited 2020 Dec 4]. Available from: https://www.ecdc.europa.eu/en/geographical-distribution-2019-ncov-cases.

6. COVID-19: infection prevention and control guidance (2020). Retrieved from https://assets.publishing.service.gov.uk/government/uploads/system/uploads/attachment_data/file/886668/COVID-19_Infection_prevention_and_control_guidance_complete.pdf6

7. Maiduguri’s keke are spreading COVID-19 prevention message (2020). Retrieved from https://reliefweb.int/report/nigeria/maiduguri-s-keke-are-spreading-covid-19-prevention-messages [Online Resource] 7

8. WaterAid Nigeria:Tackling COVID-19 in Nigeria (2020). Retrieved from https://www.wateraid.org/ng/covid-19 [Online Resource] 8

9. Thunstrom, L., Ashworth, M., Finoff, D., Newbold, S. Hesitancy towards a COVID-19 Vaccine and Prospects for Herd immunity. Available at SSRN: https://ssrn.com/abstract=3593098 or http://dx.doi.org/10.2139/ssrn.35930989

10. Coronavirus disease (COVID-19) Situation Report – 121. Data as received by WHO from national authorities by 10:00 CEST, 20 May 2020. https://www.who.int/docs/default-source/coronaviruse/situation-reports/20200520-covid-19-sitre121.pdf?sfvrsn=c4be2ec6_210

11. Piltch-Loeb, R., DiClemente R. The vaccine uptake continuum: Applying social science theory to shift vaccine hesitancy. Vaccines (Basel). 2020;8(1):76. Doi:10.3390/vaccines801007611

12. Pan American Health Organization. The immunization program in the context of the COVID-19 Pandemic. PAHO Immunization newsletter volume XLII Number 1 (March 2020).

13. Saadatian-Elah, M., Niang, I., Nabiev R., Lafond, A. et al.. Demand side interventions to increase and sustain vaccine uptake. Groupment Hospitalier Edouard Herriot. Service d’HygieLJne, EpideLJmiologie et PreLJvention. https://www.fondation-merieux.org/en/events/demand-side-interventions-increase-sustain-vaccination-uptake/

14. Adedokun, S.T., Uthman, O.A., Adekanbi, V.T. et al. Incomplete childhood immunization in Nigeria: a multilevel analysis of individual and contextual factors. BMC Public Health 17, 236 (2017). https://doi.org/10.1186/s12889-017-4137-7

15. Abdullahi, L.H., Kagina, B.M., Ndze, V.N., Hussey, G.D. Improving vaccination uptake among adolescents. Cochrane database of systematic reviews (January 2020). https://doi.org/10.1002/14651858.CD011895.pub2

16. Ogundele, O.A., Ogundele, T., Vaccine hesitancy in Nigeria: Contributing factors - way forward. The Nigerian journal of general practice Vol 18, issue 1, Page 1-4 (Jan 2020). Doi:10.4103/1008-682X.275508

17. Ochu, CL., Beynon, CM. Hepatitis B vaccination coverage, knowledge and sociodemographic determinants of uptake in high risk public safety workers in Kaduna State, Nigeria: a cross sectional survey. BMJ Open. 2017; 7:e015845. Doi: 10.1136/bmjopen-2017-015845

18. Omotowo, I.B., Meka, I.A., Ijoma, U.N. et al. Uptake of hepatitis B vaccination and its determinants among health care workers in a tertiary health facility in Enugu, South-East, Nigeria. BMC Infect Dis 18, 288 (2018). https://doi.org/10.1186/s12879-018-3191-9

19. Nigeria Population; Elaboration of data by United Nations, Department of Economic and Social Affairs, Population Division. World Population Prospects: The 2019 Revision. Available from: https://www.worldometers.info/world-population/nigeria-population/ (Accessed May 18, 2020)

20. Jekel, J., Katz, D., Elmore, J. Sample size, randomization, and probability theory. Epidemiology, Biostatistics and Preventive Medicine. 2001 2nd Ed. Philadelphia: Saunders.

21. National Demographic Health Survey (NDHS). Household population and Housing characteristics National Population Commission (NPC) Federal Republic of Nigeria, Abuja, Nigeria. 2013:11–29

22. Geldsetzer, P. Knowledge and Perceptions of COVID-19 Among the General Public in the United States and the United Kingdom: A Cross-sectional Online Survey. Annals Int Med. 2020. doi: 10.7326/M20-0912

23. Zhong B., Luo W., Li H, Zhang Q., Liu X., Li W., et al. Knowledge, attitudes, and practices towards COVID-19 among Chinese residents during the rapid rise period of the COVID-19 outbreak: a quick online cross-sectional survey. Int J Biol Sci. 2020;16(10):1745–52.

24. Abdelhafiz, A., Mohammed, Z., Ibrahim, M., Ziady, H.H., Alorabi, M., Ayyad, M., et al. Knowledge, Perceptions, and Attitude of Egyptians Towards the Novel Coronavirus Disease (COVID-19) J Community Health. 2020; 45(5):881–90.

25. Malik A., McFadden S., Etharake J., Omer S. Determinants of COVID-19 vaccine acceptance in the US. The Lancet. 2020 September; 26:100495

26. Fu, C., Wei, Z., Pei, S., Li, S., Liu, P. Acceptance and preference for COVID-19 vaccination in healthcare workers. https://doi.org/10.1101/2020.04.09.20060103.

27. Wang J., Jing R., Lai X., Zhang H., Lyu Y., Knoll M., Fang H. Acceptance of COVID-19 vaccination during the COVID-19 Pandemic in China. Vaccines 2020. 8(3), 482; https://doi.org/10.3390/vaccines8030482

28. Padhi B., Al-Mohaithef M. Determinants of intent to uptake Coronavirus vaccination among respondents in Saudi Arabia: a web-based study. [Internet]. 2020 [cited 2020 December 4]. Available from: https://www.medrxiv.org/content/10.1101/2020.05.27.20114413:

29. Lazarus J., Ratzan S., Palayew A., Gostin L., Larson H., Rabin K., et al. Global survey of potential acceptance of a COVID-19 vaccine. Nat Med (2020) October. https://doi.org/10.1038/s41591-020-1124-9.

